# Leveraging Shared Genetic Architecture Between Psychiatric and Other Medical Disorders Increases Power and Reveals Novel Psychiatric Risk Loci

**DOI:** 10.64898/2026.10.01.26364453

**Authors:** Qingyi Zhong, Apostolia Topaloudi, Peristera Paschou

## Abstract

Psychiatric disorders frequently co-occur with a broad range of non-psychiatric conditions, suggesting shared biological mechanisms that transcend traditional diagnostic boundaries. Although recent studies have demonstrated substantial genetic overlap between psychiatric and other medical disorders, the genomic architecture underlying these relationships and the extent to which shared genetic liability can be leveraged for psychiatric gene discovery remain incompletely understood. Here, we systematically investigated the shared genetic architecture between eleven major psychiatric disorders and thirty-nine non-psychiatric disorders spanning cardiovascular, metabolic, autoimmune, neurological, gastrointestinal, respiratory, and other disease categories. To maximize statistical power, we meta-analyzed genome-wide association study (GWAS) summary statistics from large biobanks and international consortia.

Genome-wide analyses identified 298 significant genetic correlations between psychiatric and non-psychiatric disorders, revealing pervasive shared liability across disease domains. Local genetic correlation analyses identified 180 genomic regions contributing to cross-disorder risk, while colocalization analyses revealed 1,470 candidate shared variants. Several highly pleiotropic loci, including regions containing FURIN and TCF4, contributed to risk across multiple psychiatric and other medical conditions. Genomic structural equation modeling identified recurrent dimensions of shared liability, including cardiovascular-kidney-metabolic and neuroimmune-related dimensions, suggesting that psychiatric-medical comorbidity is organized into biologically coherent systems.

Leveraging these shared genetic architectures through cross-trait analyses identified 1,569 significant loci, including 382 psychiatric risk loci not reported in the individual psychiatric GWAS used in our analyses. Integrative positional and functional gene-mapping approaches prioritized genes involved in neuronal development, synaptic organization, neurotrophic signaling, and cellular homeostasis, with LRFN5, MED19, PITPNM2, CTNND1, TMEM106B, GLYCTK, and PCLO as the top implicated genes. Pathway and tissue-enrichment analyses further demonstrated convergence on neurodevelopmental and synaptic processes and significant enrichment across multiple brain regions. Our results provide a comprehensive characterization of the genetic architecture underlying comorbidity between psychiatric and other medical disorders and demonstrate that leveraging genetically correlated medical conditions across domains can substantially increase power for psychiatric risk-locus discovery.

## Introduction

Psychiatric disorders frequently co-occur with a broad range of non-psychiatric conditions, including cardiovascular^1^, metabolic^2^, neurological^3^, autoimmune^4^, respiratory^5^, and gastrointestinal^6^ diseases. These disorders are also highly heritable and polygenic^7–9^, with extensive genetic overlap observed across psychiatric diagnostic boundaries^10,11^. Increasing evidence suggests that shared genetic liability extends beyond psychiatry, linking psychiatric and other medical disorders through biological mechanisms that transcend traditional disease boundaries.

Recent genetic studies support extensive overlap between psychiatric disorders and multiple domains of non-psychiatric illness. Studies of metabolic traits have revealed widespread shared genetic risk with psychiatric disorders and identified hundreds of pleiotropic variants^12^. Similar cross-domain genetic overlap has been reported for immune-mediated diseases^13^ and neurological disorders^14^. Most recently, Lawrence and colleagues examined genetic relationships between psychiatric disorders and 73 non-psychiatric illnesses spanning multiple organ systems^15^. Using genomic structural equation modeling (Genomic SEM)^16^, they demonstrated pervasive shared genetic liability between several psychiatric domains and broad systems of physical disease, identifying substantial genome-wide correlations between internalizing, neurodevelopmental, and substance-use disorders and multiple categories of medical illness. These findings provide compelling evidence that shared genetic risk extends across traditional psychiatric and medical boundaries.

Despite these advances, several important questions remain unresolved. Most studies have focused primarily on genome-wide patterns of shared liability, leaving the local genomic architecture underlying psychiatric-medical comorbidity largely unexplored. Although broad genetic correlations have been established, the specific genomic regions and pleiotropic variants responsible for these relationships remain incompletely characterized, as does the higher-order structure of shared liability across medical conditions. Most importantly, it remains unclear whether shared genetic architecture across psychiatric and medical disorders can be leveraged to improve psychiatric locus and gene discovery. Addressing these questions requires moving beyond the identification of genetic correlations to characterize the genomic architecture and organization of cross-domain sharing and determine whether this information can be exploited for psychiatric genetic discovery.

Here, we systematically investigated the shared genetic architecture between eleven major psychiatric disorders and thirty-nine non-psychiatric disorders spanning multiple disease categories. We expanded available GWAS through meta-analysis and integrated genome-wide and local genetic correlation analyses, colocalization, and Genomic SEM to characterize shared liability across disorders and identify pleiotropic genomic mechanisms underlying psychiatric-medical comorbidity. We then leveraged these shared genetic architectures in cross-trait analyses to increase power for psychiatric risk-locus discovery, followed by integrative functional analyses to identify implicated genes and biological pathways. This framework resolves the genomic architecture underlying psychiatric-medical comorbidity and demonstrates its utility for psychiatric gene discovery.

## Results

An overview of the study design and analytical workflow is shown in Figure 1.

**Figure 1.**
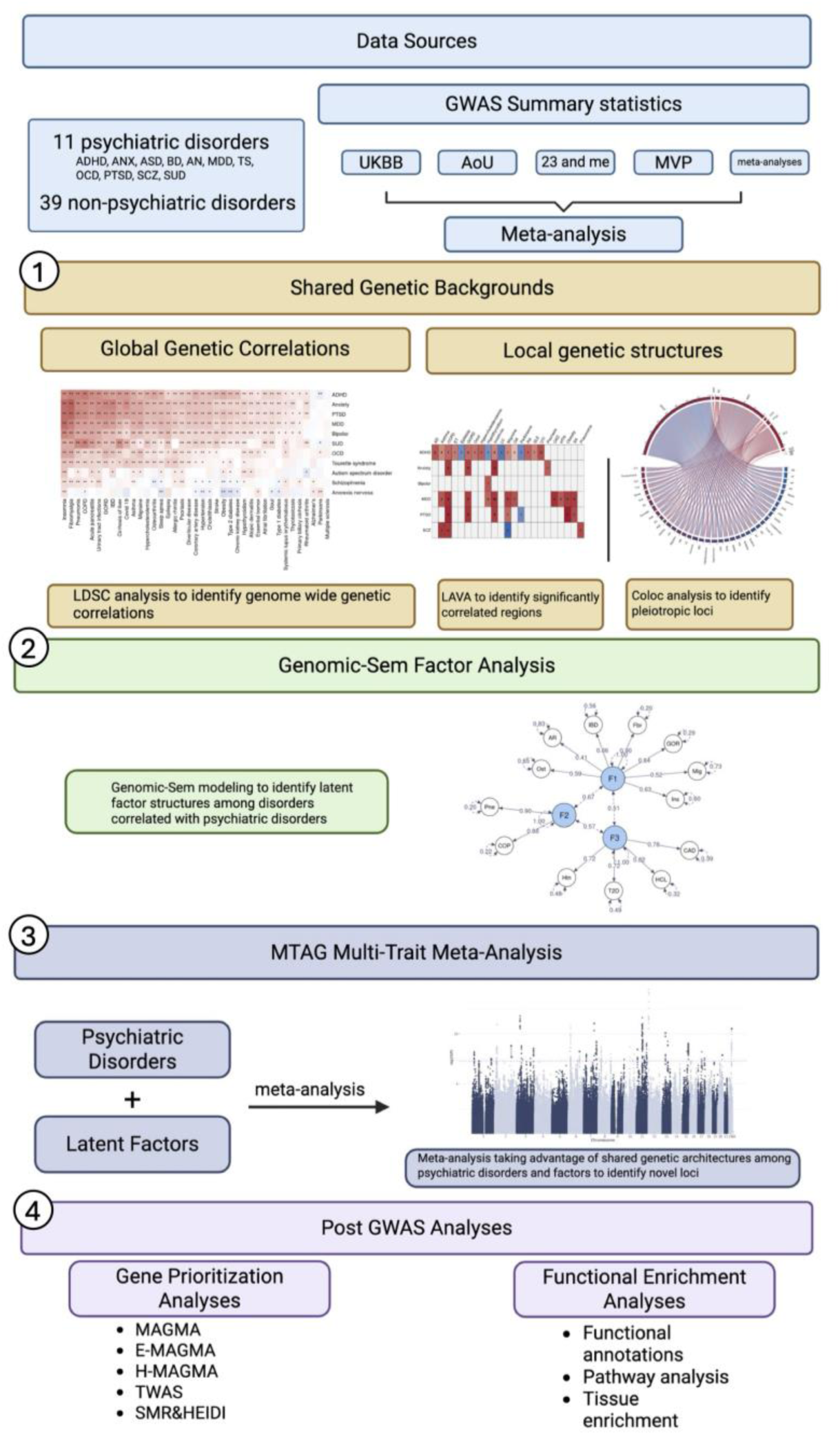
Analytical framework for identifying shared genetic architecture and novel psychiatric risk loci. GWAS summary statistics from 11 psychiatric disorders and 39 non-psychiatric disorders were meta-analyzed across large biobanks and international consortia. Genome-wide and local genetic analyses were used to identify shared liability, pleiotropic genomic regions, and candidate causal variants linking psychiatric and non-psychiatric disorders. Genomic SEM was applied to derive latent factors representing shared medical risk dimensions. Leveraging these shared genetic architectures, cross-trait analyses were performed to identify novel psychiatric risk loci. Subsequent gene-prioritization, pathway-enrichment, and tissue-enrichment analyses were used to characterize the biological mechanisms underlying psychiatric-medical comorbidity.

To maximize statistical power and harmonize available genetic data, we expanded GWAS meta-analyses for psychiatric and non-psychiatric disorders integrating summary statistics from large biobanks and international consortia (Methods and Supplementary Tables 1-3). Notably, several meta-analyses substantially increased sample sizes relative to the largest published studies, with the greatest gains observed for migraine, hypertension, and GORD (1,654,231, 1,013,710, and 1,010,029 additional individuals, respectively), thereby increasing power for downstream cross-trait analyses. Supplementary Table 4 provides a comprehensive summary of the cases, controls, total sample size, heritability, as well as the increase in sample size compared with the largest published study. All disorders demonstrated robust heritability estimates and limited evidence of confounding, supporting their suitability for downstream cross-disorder analyses.

### Pervasive shared genetic liability links psychiatric and non-psychiatric disorders

To systematically characterize shared genetic liability across disease domains, we estimated genome-wide genetic correlations between eleven psychiatric disorders and thirty-nine non-psychiatric disorders, covering a broad range of disorder categories including cardiovascular, metabolic, respiratory, neurological, gastrointestinal, immune and musculoskeletal disorders. We identified 298 pairs of disorders across domains to be significantly correlated, revealing extensive genetic overlap between psychiatric disorders and a broad range of other medical conditions (Figure 2; Supplementary Table 5).

**Figure 2.**
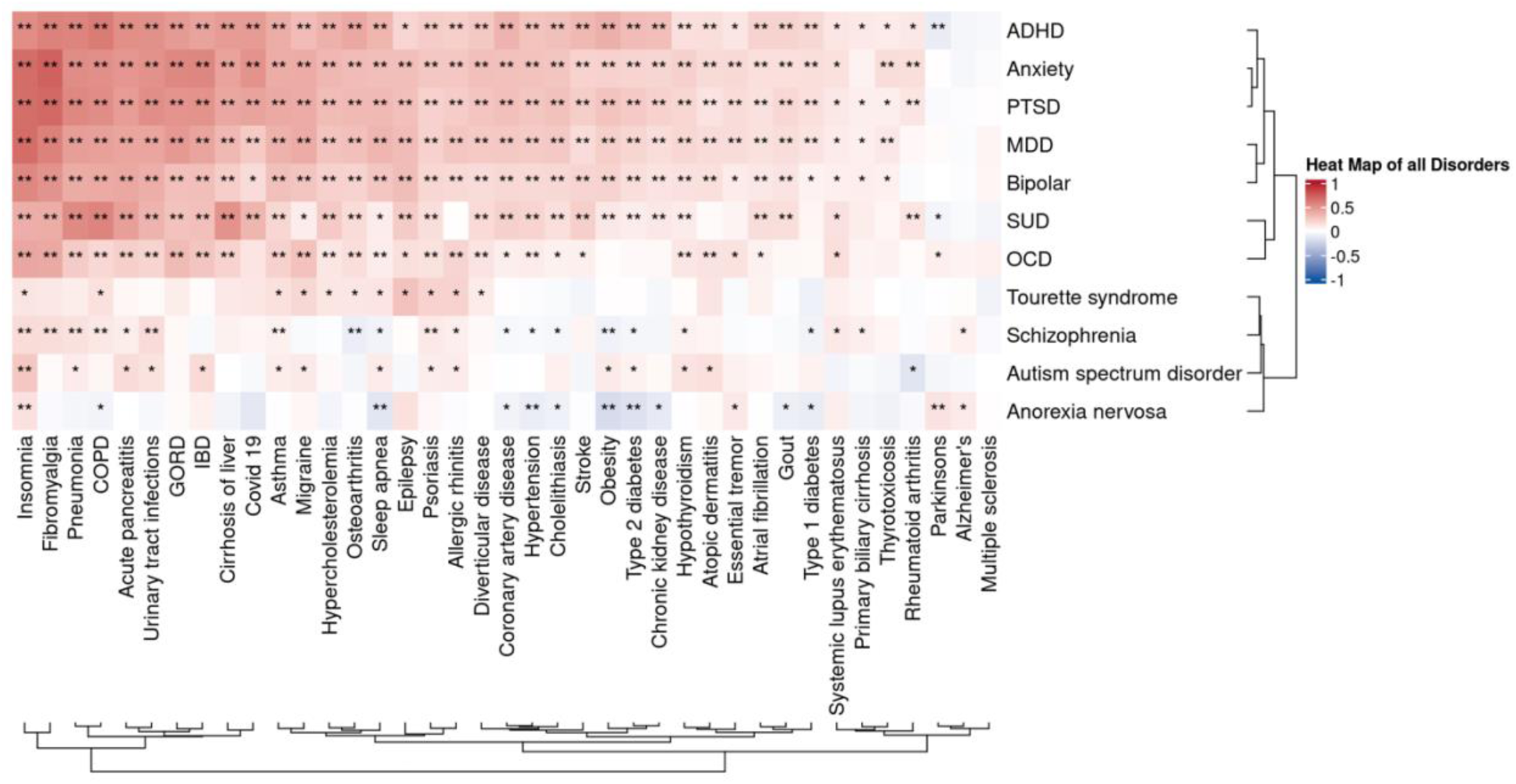
Extensive shared genetic liability between psychiatric and non-psychiatric disorders. Heatmap of genome-wide genetic correlations (rg) between 11 psychiatric disorders and 39 non-psychiatric disorders estimated using linkage disequilibrium score regression (LDSC). Colors represent the direction and magnitude of genetic correlation, with red indicating positive correlations and blue indicating negative correlations. Asterisks indicate significant associations after multiple-testing correction (*FDR-adjusted P < 0.05; **Bonferroni-adjusted P < 0.05). The widespread positive correlations highlight extensive shared genetic liability across disease domains. Exact estimates are provided in Supplementary Table S5.

ADHD exhibited the broadest pattern of shared genetic liability, showing significant correlations with 37 non-psychiatric disorders. Among non-psychiatric medical conditions, insomnia emerged as the trait most consistently associated across clinical domains, displaying significant positive genetic correlations with all eleven psychiatric disorders examined. The strongest correlations were observed between anxiety and fibromyalgia (rg = 0.73, SE = 0.05), PTSD and fibromyalgia (rg = 0.70, SE = 0.04), and between insomnia and several psychiatric disorders, including anxiety (rg = 0.69, se = 0.03), MDD (rg = 0.70, se = 0.02), and PTSD (rg = 0.69, se = 0.02). Collectively, these findings indicate that shared genetic liability between psychiatric and medical disorders is widespread rather than confined to specific disease categories.

### Shared genetic liability localizes to specific genomic regions

Although genome-wide correlations demonstrate broad genetic overlap, they do not identify the genomic regions responsible for these relationships. We therefore performed local genetic correlation analyses to map shared liability across the genome. Across 298 correlated disorder pairs, we identified 180 genomic regions exhibiting significant local genetic correlations (Figure 3; Supplementary Table 6). ADHD showed local genetic sharing with the broadest range of non-psychiatric disorders, whereas MDD displayed the largest total number of significantly correlated genomic regions, with particularly extensive local sharing with insomnia. Most local correlations were positive and mirrored the patterns observed at the genome-wide level.

**Figure 3.**
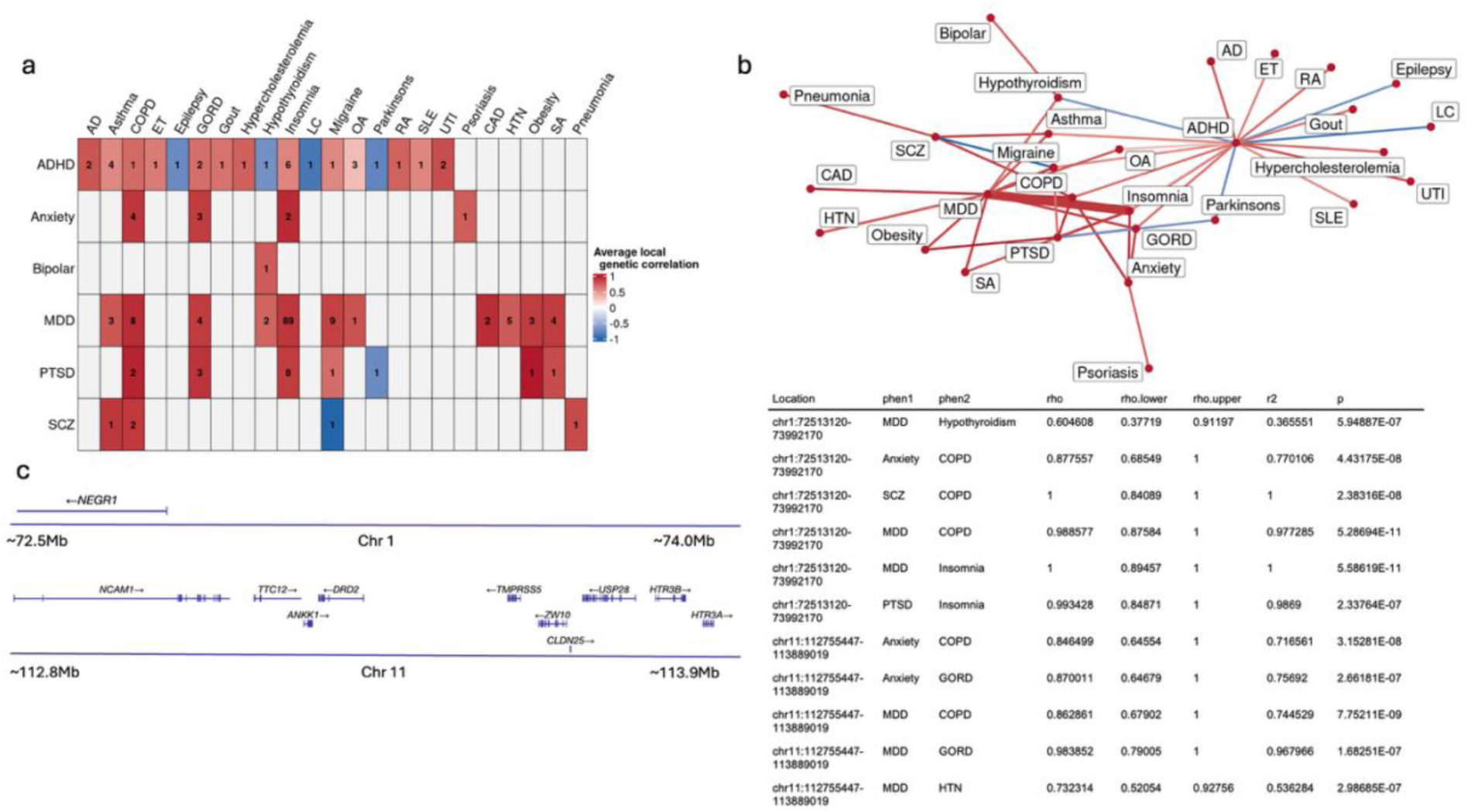
Shared genetic liability localizes to specific genomic regions. **a,** Heatmap of significant local genetic correlations between psychiatric and non-psychiatric disorders identified using LAVA. Colors indicate the mean local genetic correlation across significant loci. **b,** Network of local genetic sharing between disorders. Edge width represents the number of significant loci shared between disorder pairs, and edge color represents the average local genetic correlation. **c,** Representative pleiotropic loci on chromosomes 1 and 11 showing extensive local genetic sharing across multiple psychiatric and non-psychiatric disorders. Gene annotations are displayed for each region. The table summarizes local genetic correlation estimates (ρ), 95% confidence intervals, variance explained (R²), and significance values. (Full results are shown in Supplementary Table S6)

Nine loci emerged as hotspots of cross-disorder genetic sharing (significant among at least 3 different pairs of disorders). The strongest signal was observed at locus chr1:72513120-73992170 which showed significant correlations in 6 disorder pairs. This locus contains NEGR1, a gene involved in neuronal development, cell adhesion, and synapse formation. Another prominent region at chr11:112755447-113889019 was shared among 5 disorder pairs, and it encompassed the NCAM1–TTC12–ANKK1–DRD2 gene cluster, a locus previously implicated in psychiatric disorders and dopaminergic signaling. These findings demonstrate that shared genetic liability between psychiatric and other medical disorders is concentrated, in part, within discrete genomic regions harboring biologically relevant risk genes.

### Colocalization analyses identify widespread pleiotropic variants

To determine whether shared regional associations reflected common underlying potentially causal variants, we performed Bayesian colocalization analyses across correlated psychiatric and non-psychiatric disorder pairs. We identified 1,470 candidate pleiotropic variants with strong support for a shared causal signal (PP.H4 ≥ 0.8; Figure 4). ADHD exhibited the greatest degree of variant sharing with non-psychiatric disorders, particularly asthma. Notably, 635 variants were shared across multiple disorder pairs, indicating extensive pleiotropy across disease domains. (Supplementary Table S7) The most broadly shared variant, rs4702 within FURIN, was implicated in 138 psychiatric and non-psychiatric disorder pairs involving 6 psychiatric disorders. Given the established role of FURIN in neurodevelopmental and signaling pathways, these findings provide direct evidence that common genetic variants contribute to liability across both psychiatric and other medical disorders.

**Figure 4.**
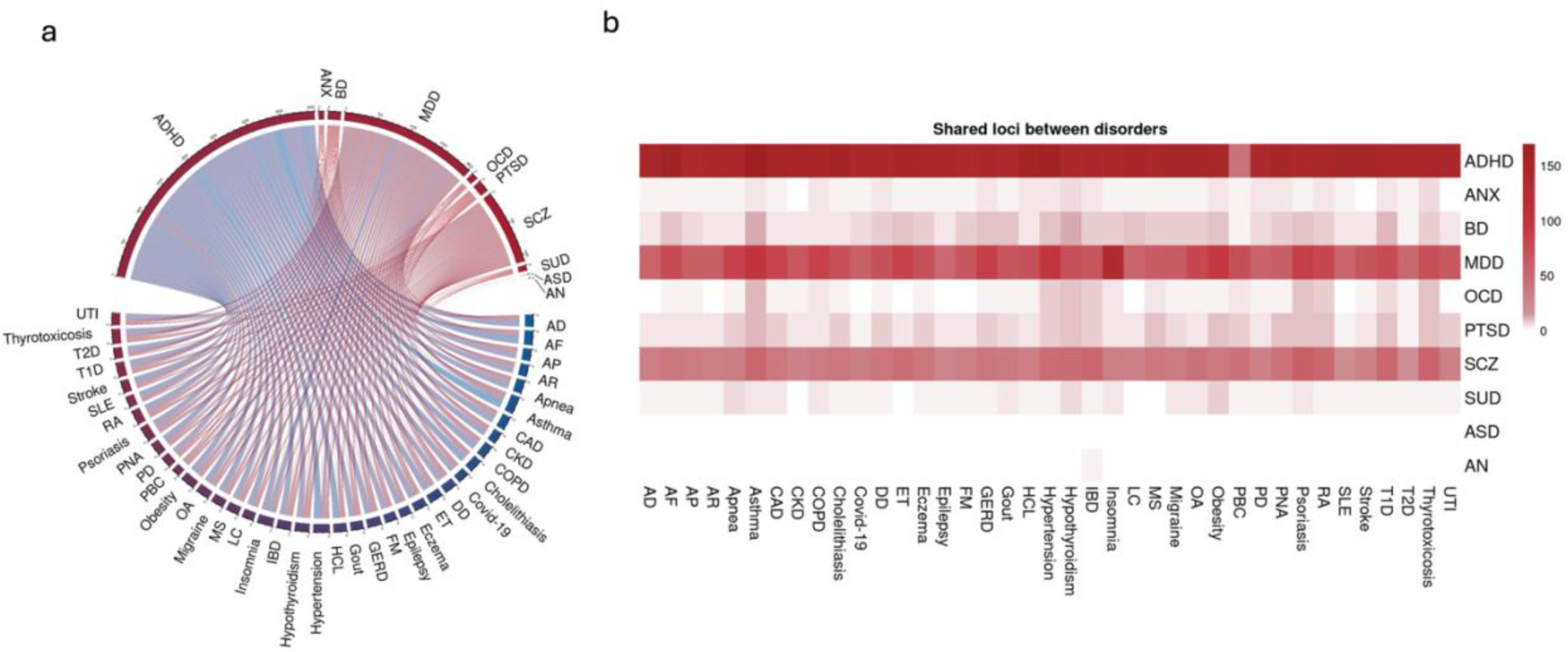
Colocalization analyses reveal widespread pleiotropic variants linking psychiatric and non-psychiatric disorders. **a,** Chord diagram illustrating the number of shared candidate causal variants between disorder pairs identified through colocalization analyses. Ribbon width is proportional to the number of shared variants, highlighting major sources of cross-disorder genetic overlap. **b,** Heatmap showing pairwise sharing of candidate causal variants across psychiatric and non-psychiatric disorders. Color intensity reflects the number of shared variants. These results demonstrate extensive pleiotropic genetic effects underlying psychiatric-medical comorbidity. The complete set of variants is reported in Supplementary Table S7.

### Latent factors capture major dimensions of shared psychiatric-medical risk

To better characterize higher-order patterns of shared genetic liability, we applied Genomic SEM to non-psychiatric disorders genetically correlated with each psychiatric disorder. Across most psychiatric disorders, two- and three-factor models provided the best fit to the data (Figure 5; Supplementary Table 8).

**Figure 5.**
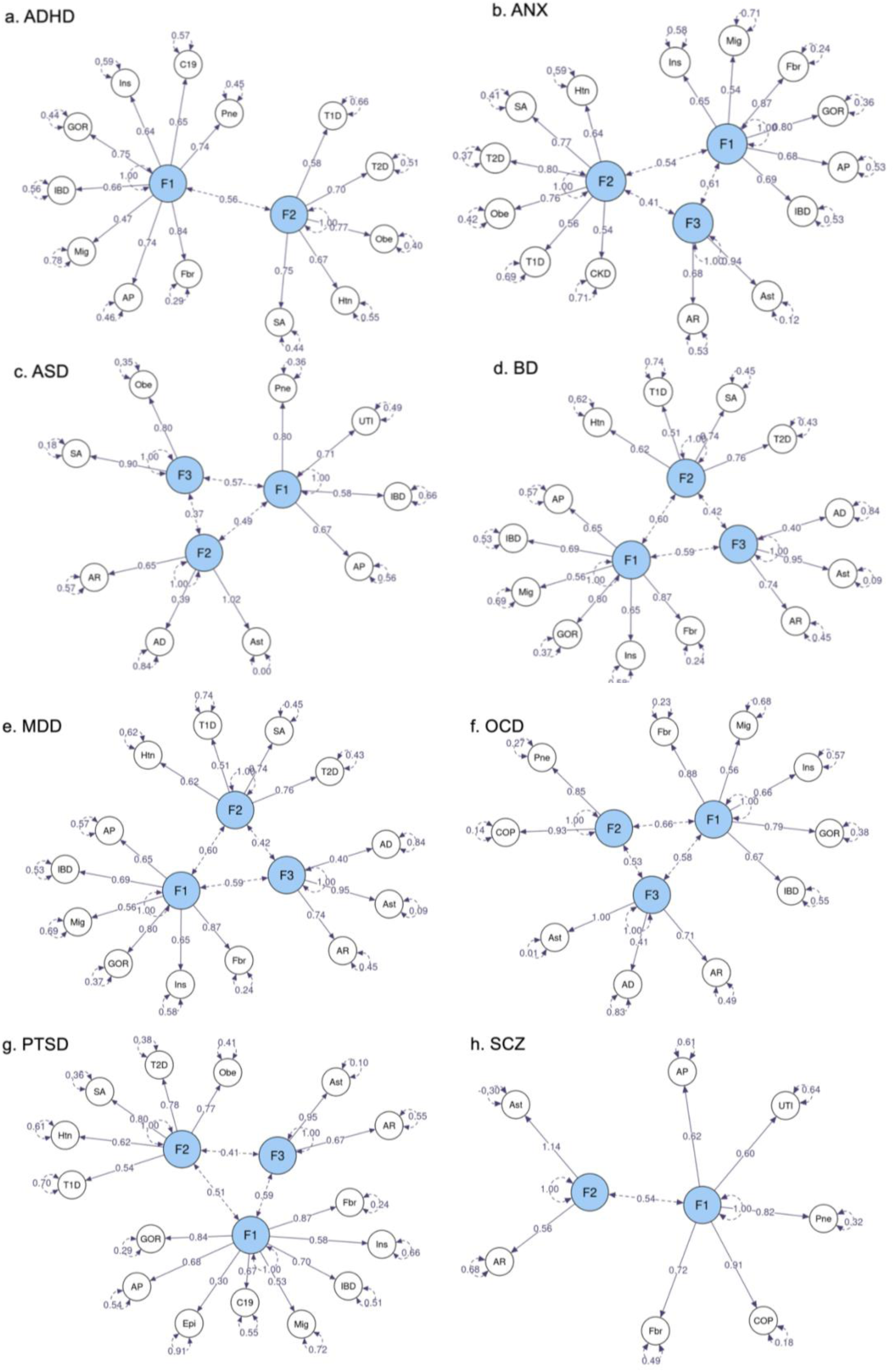
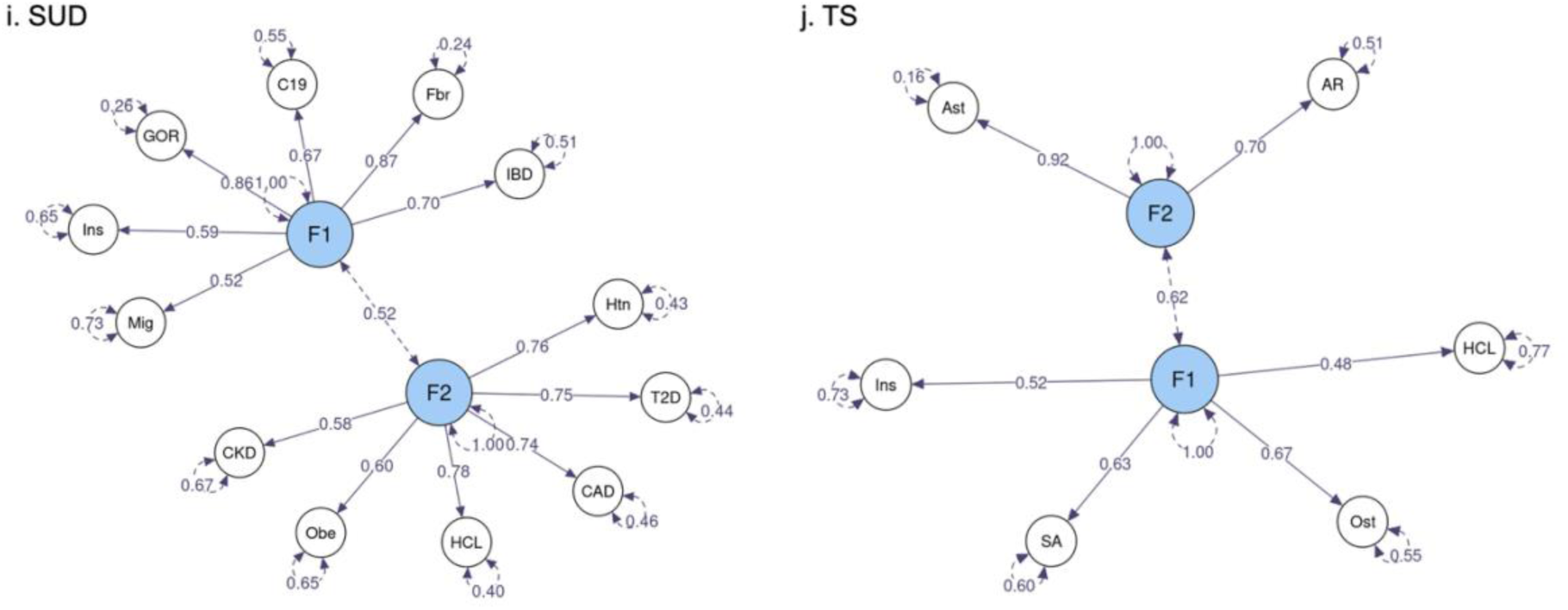
Shared genetic liability between non-psychiatric disorders is organized into latent genomic dimensions. Path diagrams of the best-fitting Genomic SEM models for non-psychiatric disorders genetically correlated with each psychiatric disorder. Latent factors capture shared genetic variance across correlated medical conditions. Single-headed arrows indicate standardized factor loadings, double-headed arrows between factors indicate inter-factor genetic correlations, and double-headed arrows connected to disorders indicate residual genetic variance. Standardized estimates are shown for all paths. Panels correspond to **a,** ADHD; **b,** ANX; **c,** ASD; **d,** BD; **e,** MDD; **f,** OCD; **g,** PTSD; **h,** SCZ; **i,** SUD; and **j,** TS.

Despite differences in factor structure across psychiatric disorders, recurring patterns of medical trait loadings revealed several common dimensions of shared liability. One prominent factor encompassed cardiovascular, metabolic, and renal conditions, including hypertension, coronary artery disease, type 2 diabetes, obesity, stroke, and chronic kidney disease. A second factor was characterized by neuroimmune and inflammatory conditions, including allergic rhinitis, fibromyalgia, gastroesophageal reflux disease, inflammatory bowel disease, migraine, and insomnia. Inter-factor correlations ranged from 0.37 to 0.66, indicating substantial overlap among these dimensions of medical risk. These results suggest that psychiatric disorders share genetic liability with coherent biological systems rather than with isolated medical conditions.

### Leveraging shared genetic architecture reveals novel psychiatric risk loci

Having characterized the extent and structure of shared genetic liability across psychiatric and non-psychiatric disorders, we next asked whether this shared architecture could be leveraged to increase power for psychiatric risk-locus discovery. Multivariate GWAS of the latent factors identified variants associated with shared dimensions of medical risk while excluding loci exhibiting significant heterogeneity across constituent disorders. All latent factors remained significantly genetically correlated with their corresponding psychiatric disorders (Supplementary Table 9), supporting their relevance as genetically informed cross-disorder traits.

We then performed MTAG analyses integrating each psychiatric disorder with the genetically correlated latent medical factors identified for that disorder. Cross-trait analyses identified 1,569 significant loci and increased power for psychiatric risk-locus discovery, identifying 382 loci that did not overlap with loci previously reported in the individual psychiatric GWAS used in this analysis (Supplementary Tables 10–11). MTAG increased the GWAS-equivalent sample size across most psychiatric disorders, with the greatest increases observed for SUD (584,584) and MDD (360,384) (Supplementary Table 12). Functional annotation indicated that significant variants were predominantly noncoding, with most mapping to intergenic or intronic regions (Supplementary Table 13).

Several newly identified variants demonstrated effects across multiple psychiatric disorders. The most significant associations included variants in DPP6, NLGN1, CASC10, SKIDA1, and MAML3, implicating pathways involved in neuronal excitability, synapse organization and transcription regulation. Together, these findings demonstrate that leveraging shared genetic architecture across disease domains substantially increases power for psychiatric gene discovery.

To further characterize the genes underlying these associations, we performed MAGMA gene-based analyses using the multi-trait GWAS results. MAGMA identified 953 genes reaching Bonferroni-corrected significance across the psychiatric disorders. DCC emerged as the most pleiotropic gene significant across 8 psychiatric disorders. DCC encodes the netrin-1 receptor, a key regulator of axon guidance and neuronal circuit formation during neurodevelopment.

To place the findings into a biological context, we performed pathway and tissue-enrichment analyses using MAGMA (Supplementary Table 14). Across psychiatric disorders, results showed consistent enrichment for pathways related to synaptic structure, synaptic membrane function, postsynaptic specialization, neuron generation, and neurogenesis (Table 1, Supplementary Table 15). Tissue-enrichment analyses further demonstrated significant enrichment in brain and pituitary tissues, with multiple specific brain regions showing robust associations (Table 1 and Supplementary Tables 16-17). The top enriched pathway and brain region for each disorder are provided in Table 1.

**Table 1.**
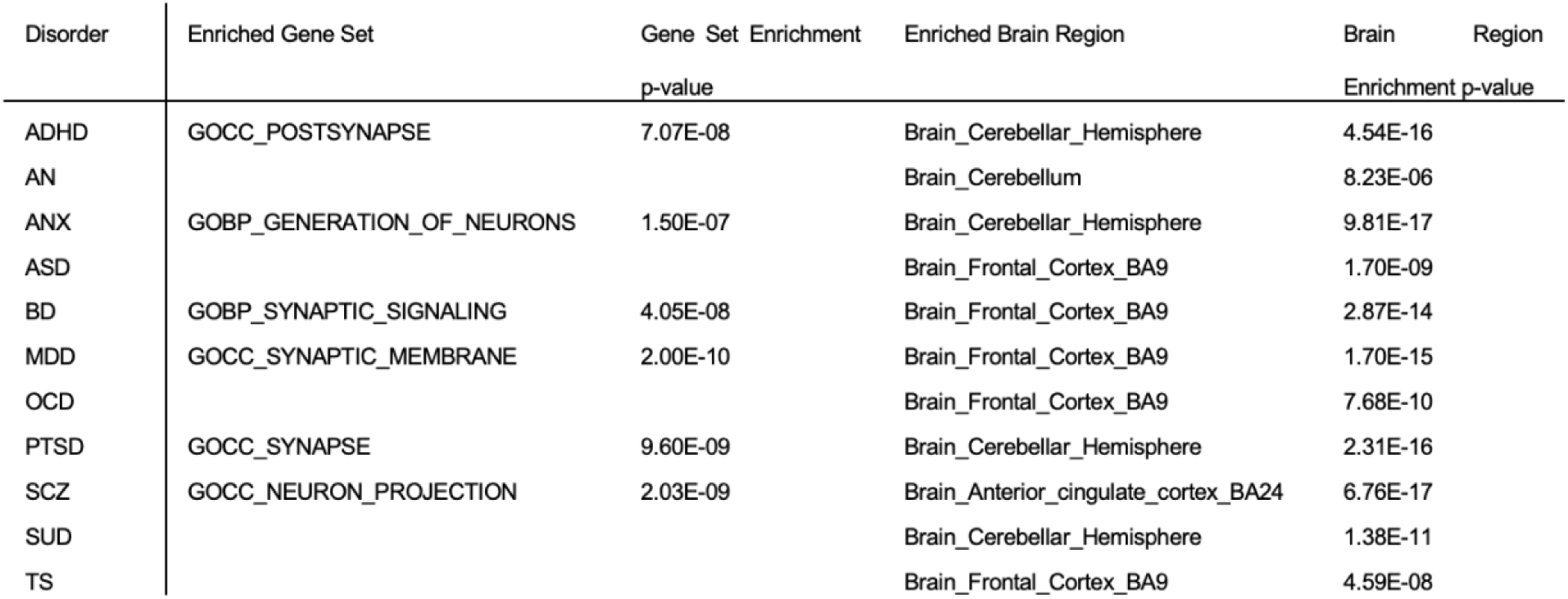
Most significantly enriched pathway and brain region for each psychiatric disorder.

### Convergent gene prioritization implicates neuronal development, synaptic function, and proteostasis

We further integrated evidence across multiple complementary functionally informed gene-prioritization approaches, including e-MAGMA, h-MAGMA, TWAS, and SMR analyses (Supplementary Tables 18-21).

Across all methods, 3,873 unique genes were significantly associated with at least one psychiatric disorder. Applying a more stringent criterion requiring support from at least three independent approaches yielded 428 high-confidence genes. 115 of these genes showed associations across multiple psychiatric disorders, with PITPNM2, CTNND1, TMEM106B, GLYCTK, and PCLO each associated with 6 disorders (Supplementary Table 22). Importantly, comparison with genes reported in previous GWAS from the GWAS Catalogue identified 3,351 novel genes, including 252 high-confidence candidates supported by multiple lines of evidence.

These findings indicate that shared genetic liability across psychiatric and medical disorders converges on core neurodevelopmental and synaptic processes, providing a biological framework linking pleiotropic genetic risk to psychiatric disease susceptibility.

## Discussion

Recent genomic studies have established substantial genetic overlap between psychiatric disorders and other medical conditions, but the architecture underlying this overlap and its potential utility for psychiatric gene discovery remain incompletely understood. Here, we show that shared psychiatric-medical liability extends from genome-wide correlations to specific genomic regions and shared genetic signals and is organized into recurrent dimensions spanning major systems of medical disease. Importantly, leveraging this shared architecture substantially increased power for psychiatric risk-locus discovery. These findings move beyond documenting genetic overlap across disease boundaries to demonstrate how its underlying structure can inform psychiatric gene discovery.

A central finding of our study is the remarkable extent of shared genetic liability across disease categories. We identified 298 significant genetic correlations between psychiatric and non-psychiatric disorders, indicating that shared genetic risk extends far beyond traditional psychiatric boundaries. ADHD exhibited the broadest pattern of overlap with medical conditions, while insomnia emerged as one of the most widely associated non-psychiatric disorders, displaying significant correlations across all eleven psychiatric disorders examined. These findings are consistent with extensive epidemiological evidence demonstrating elevated rates of cardiovascular, metabolic, neurological, autoimmune, and gastrointestinal disorders among individuals with psychiatric conditions^1–6^. They also extend previous genomic studies focused on metabolic^17^, immune^13^, and neurological^18^ disorders, as well as recent work of Lawrence and colleagues^15^ demonstrating widespread shared genetic liability between psychiatric domains and systems of physical disease. Here, we build on this work by resolving the local genomic architecture underlying these relationships and identifying the specific regions and shared genetic signals contributing to cross-domain liability.

While genome-wide correlations establish the existence of shared liability, they do not identify the genomic mechanisms responsible for these relationships. By integrating local genetic correlation and colocalization analyses, we demonstrate that psychiatric-medical comorbidity is concentrated within specific genomic regions and shared variants. We identified 180 significant local genetic correlations and 1,470 candidate pleiotropic variants, indicating that shared liability is driven by discrete genomic loci rather than diffuse genome-wide effects alone. Several loci emerged as prominent hubs of cross-disorder sharing. Among these, the FURIN locus was implicated across an exceptionally large number of disorder pairs. FURIN encodes a proprotein convertase involved in neuronal signaling and neurodevelopment and has previously been implicated in schizophrenia and related psychiatric phenotypes^19^. Experimental studies have demonstrated that FURIN perturbation influences neurite growth and neuronal activity, providing biological support for its role as a pleiotropic mediator of disease risk^20^. Similarly, variants within TCF4 were associated with a broad range of psychiatric and non-psychiatric conditions. TCF4 plays a critical role in inhibitory interneuron development^21^ and has previously been implicated in ASD, MDD, PTSD, bipolar disorder, and schizophrenia^22^. Associations with cardiovascular and renal phenotypes^23,24^ further suggest that TCF4 may represent a biologically important link between psychiatric and somatic disease processes. These findings suggest that a subset of highly pleiotropic loci may contribute disproportionately to shared risk across multiple disease domains.

Our factor analyses further revealed that shared liability is organized into biologically meaningful dimensions. Despite differences in factor structure across psychiatric disorders, recurring patterns emerged, including a cardiovascular-kidney-metabolic dimension and a second dimension encompassing neurological, pain, sleep, gastrointestinal, and inflammatory phenotypes. The cardiovascular-kidney-metabolic factor included chronic kidney disease, obesity, hypertension, coronary artery disease, diabetes, hypercholesterolemia, and sleep apnea, closely mirroring the recently proposed cardiovascular-kidney-metabolic (CKM) syndrome framework^25^. This convergence suggests that common genetic influences may underlie the interconnected pathophysiology of metabolic, cardiovascular, and renal disorders. Likewise, the neuroimmune-related factor incorporated insomnia, migraine, fibromyalgia, gastroesophageal reflux disease, and epilepsy. Previous clinical and epidemiological studies have documented substantial overlap among these conditions^26,27^, and our results provide evidence that such relationships are partly attributable to shared genetic architecture. More broadly, these findings indicate that psychiatric disorders share liability with coherent biological systems rather than with isolated medical conditions, supporting the utility of dimensional approaches for understanding disease risk.

Perhaps one of the most important findings of our study is that shared genetic architecture can be leveraged to improve psychiatric gene discovery. By integrating psychiatric disorders with genetically correlated medical factors, we identified 1,569 significant loci, including 382 psychiatric risk loci not reported in the individual psychiatric GWAS used in our analyses. While previous studies have demonstrated shared liability between psychiatric and non-psychiatric disorders ^13–15^, our results extend this work by showing that genetically correlated medical phenotypes contain information that can increase power to identify psychiatric associations. More broadly, these findings demonstrate the value of leveraging genetic relationships across conventional disease boundaries to uncover biological mechanisms underlying complex psychiatric phenotypes.

Integration of positional and functional gene-mapping approaches implicated a large number of genes involved in neuronal development, synaptic connectivity, neurotrophic signaling, and cellular homeostasis. Several genes emerged consistently across multiple analytical approaches and psychiatric disorders. LRFN5, one of the most broadly implicated genes, plays an important role in neuronal development and synaptic membrane adhesion and was associated with ADHD, MDD, OCD, anxiety, PTSD, and substance use disorder. Previous studies have linked LRFN5 to ASD^28^, MDD^29^, and several cancers^30^, suggesting that its biological functions may extend beyond the nervous system. Likewise, MED19, PITPNM2, CTNND1, TMEM106B, GLYCTK, and PCLO were implicated across multiple disorders and analytical methods. These genes converge on biological processes related to neuronal gene expression^31^, synapse development^32^, neuronal trafficking^33^, neurotransmitter metabolism^34^, and synaptic organization^35^, providing evidence that disturbances in neuronal network formation and maintenance represent fundamental mechanisms contributing to psychiatric risk.

Our analyses also identified several potentially novel candidate genes. MAP3K9 emerged as a candidate contributor to mood and anxiety disorders, extending previous evidence implicating MAPK signaling in major depression^36^. EDEM3 was associated with ADHD and may implicate endoplasmic reticulum proteostasis and protein-quality-control pathways in disease susceptibility. CTSF, previously linked primarily to bipolar disorder^37^, showed evidence of broader involvement in mood-disorder biology. DIAPH3, a regulator of actin cytoskeletal dynamics, neurite outgrowth, and synaptic development, was associated with both anxiety and depression, suggesting a role for cytoskeletal remodeling and neuronal connectivity in psychiatric disease. Although additional functional studies will be required to establish causal relationships, these findings provide promising targets for future mechanistic investigation.

Notably, despite leveraging genetic information from a broad range of systemic medical phenotypes, the resulting associations retained a strong neurobiological signature, with enrichment for neurodevelopmental and synaptic processes and predominant expression in brain tissues. Risk genes were consistently enriched for pathways related to neuronal development, neurite growth, synapse assembly, postsynaptic organization, and synaptic signaling. These findings are highly consistent with previous GWAS and cross-disorder studies implicating neurodevelopmental and synaptic mechanisms as central contributors to psychiatric disease^15,16,38^. Additional enrichment of pathways related to transcriptional regulation, RNA binding, and ion-channel activity suggests that broader regulatory processes may also contribute to disease susceptibility^39,40^. Consistent with these observations, tissue-enrichment analyses demonstrated robust enrichment across multiple brain regions, including cortical, limbic, basal ganglia, cerebellar, and brainstem structures, with additional enrichment observed in the pituitary gland.

Several limitations should be considered when interpreting these findings. First, our analyses were restricted to individuals of predominantly European ancestry due to the availability of sufficiently powered GWAS datasets, and therefore the generalizability of these findings to other populations remains uncertain. Expanding cross-disorder analyses to more diverse ancestral groups will be an important priority for future research. Second, as is typical in studies integrating multiple cohorts, differences in phenotype definition and ascertainment across cohorts and biobanks may contribute to heterogeneity among the GWAS datasets analyzed. Genetic correlation reflects shared genetic liability but does not establish a causal relationship between psychiatric and other medical disorders, and colocalization provides statistical support for shared genetic signals rather than direct evidence of causal variants. Similarly, Genomic SEM captures shared variation represented by common variants included in GWAS datasets, and the biological interpretation of inferred latent factors will require additional validation. Multivariate approaches can increase discovery power by leveraging shared effects across traits, but newly identified loci will require replication in independent datasets and functional validation. Finally, despite substantial increases in sample size achieved through meta-analysis, statistical power remained limited for some disorders, including Tourette syndrome and anorexia nervosa, potentially reducing our ability to detect additional shared loci and biological pathways.

In summary, our findings demonstrate that shared genetic liability between psychiatric and other medical disorders is extensive, locally structured, and organized across recurrent dimensions of medical risk. Importantly, this shared architecture can be leveraged to increase power for psychiatric risk-locus and gene discovery. Cross-domain genetic sharing therefore represents not only a feature of psychiatric-medical comorbidity, but also a resource for psychiatric genetic discovery and for identifying biological mechanisms that transcend conventional disease boundaries.

## Methods

### Data selection and meta-analysis

All analyses were conducted using previously collected GWAS summary statistics obtained from publicly available resources or through approved data-access procedures. The original studies obtained approval from the relevant institutional review boards or ethics committees and informed consent from participants, as applicable. The present study involved secondary analysis of summary-level data and did not involve recruitment of participants or collection of new individual-level data.

The psychiatric disorders we focused on in this study are the eleven psychiatric disorders studied by the Psychiatric Genomics Consortium (PGC), representing the broad landscape of major psychiatric conditions, including attention deficit hyperactivity disorder (ADHD), anxiety disorder (ANX), autism spectrum disorder (ASD), bipolar disorder (BD), anorexia nervosa (AN), major depressive disorder (MDD), Tourette syndrome (TS), obsessive-compulsive disorder (OCD), post-traumatic stress disorder (PTSD), schizophrenia (SCZ), and substance use disorder (SUD). The summary statistics for the eleven disorders are publicly available and were downloaded from the PGC website^41–51^ (Table S1).

To identify the non-psychiatric disorders to include, we looked at large clinically relevant disease categories including autoimmune, cardiovascular, degenerative, gastrointestinal, infectious, metabolic, neurological, and respiratory disorders. The summary statistics of European ancestry were sourced from previous studies, consortiums, and large biobanks including Finngen^52^ (DF12), UKBB^53^, All of Us^54^, 23andMe Research Institute, and Million Veterans Program (MVP)^55^ (Table S2). For each category, disorders with large publicly available GWAS summary statistics of more than 3000 cases were included. All GWAS summary statistics were harmonized to the human genome build 37 (GRCh37/hg19) and rare variants with minor allele frequency less than 0.1% were excluded from the analysis. To ensure sufficient statistical power and robust genetic signal, only traits with heritability Z-scores larger than 3 were retained. Our final selection of disorders contained 39 disorders (Table S1-S2).

Meta-analyses were performed for each of the traits with multiple available summary statistics to increase the sample size and improve the statistical power. The meta-analyses were performed using the METAL^56^ software with the inverse variance weighted (IVW) fixed-effects method. Our meta-analyses expanded the sample sizes of several of the GWAS beyond the published data.

### Genome-wide genetic correlation

To identify disorders that were significantly correlated to psychiatric disorders, genome-wide genetic correlations between pairs of psychiatric disorders and non-psychiatric disorders were calculated using linkage disequilibrium score regression (LDSC)^57^ implemented through Genomic SEM^16^. In the analyses we included only the HapMap3 SNPs and the 1000 Genomes Project European LD reference. For the calculations we used the population prevalence values reported in previous literature (Table S3), and the sample prevalence was set to 0.5 as the effective sample size was used. The significantly correlated pairs were identified with False Discovery Rate (FDR) corrected P values less than 0.05. Downstream analyses were restricted to traits with significant positive genome-wide genetic correlations, allowing us to focus on comorbid conditions that may increase psychiatric disorder risk through shared genetic liability.

### Local genetic correlation

Since local genetic correlations may be masked in the genome-wide genetic correlation analysis, we implemented LAVA^58^ to identify regions with shared genetic association between phenotypes. We partitioned the genome into 2495 approximately equal sized and LD-independent loci as provided by LAVA’s developers, and loci with sufficient genetic signal identified using the univariate test with significant Bonferroni adjusted p value (p < 2×10^−5^) were included in the bivariate analysis for genetic correlations between trait pairs. We performed a total of 122,44 bivariate between psychiatric and non-psychiatric disorders, and considered as significant the correlations with a Bonferroni corrected p-value threshold of p<4×10^−6^ (0.05/12,244).

### Colocalization analysis

Then we used the R package Coloc^59^ to perform Bayesian colocalization analysis on the significant loci from the psychiatric disorders to determine if they were shared with the correlated somatic disorders. We identified the independent lead SNPs and risk loci using Plink^60^ clumping with p value threshold 5 × 10^−8^ and r^2^ < 0.1 within 3000kb region. Next, we implemented the Coloc.abf function to analyze the shared loci between two datasets and calculate the posterior probabilities (PP) indicating that there is no association, one causal SNP for one of or both traits, and two different causal SNPs for two traits in each locus. The posterior probability for hypothesis H4 gives the probability of the locus being pleiotropic for the traits analyzed, thus loci with PP H4 > 0.8 were considered pleiotropic.

### Genomic-SEM

#### Genome-wide models

To model the shared genetic architecture of the somatic disorders with positive genetic correlations with each psychiatric trait and construct factor models, we implemented Genomic SEM^16^. We prepared the data as described in global genetic correlation tests before inputting to the multivariable LDSC analyses. The exploratory factor analysis was performed using the factanal function from the R package stats with promax rotations. Different factor structures were tested in the exploratory factor analysis, and the correlated factor model with varying numbers of factors for different disorders was selected based on the proportion of variance explained and the interpretability of the factor components.

Confirmatory factor analysis was performed using the results from the EFA. The disorders with loadings >0.5 in each factor were included in the CFA to ensure strong contributions to the factors and avoid weak indicators that could destabilize model estimation. The residual variances were restricted to be above 0 to prevent Heywood cases. The model fit was evaluated using the CFI and SRMR. CFI measures how well the proposed model fits the actual data, and SRMR measures the difference between the observed and the model implied covariance matrix and models with CFI > 0.9 and SRMR < 0.08 were considered good fits.

#### Multivariate GWAS

To calculate the SNP associations with each of the correlated latent factors, we performed multivariate GWAS analysis using Genomic SEM. We used the sumstats function in Genomic-SEM to align the effects to the same reference allele and standardize the effect sizes and SEs relative to the unit-variance scaled phenotypes. Only the SNPs that were present in all analyzed datasets were preserved in the final combined summary statistics file. The multivariate GWAS was then conducted using the userGWAS function to estimate the association of the SNPs with the factors using the correlated factors model. The effective sample size of each factor was calculated using the formula provided in Mallard et al.^39^. We also calculated the Q-SNP p-value to assess SNP-level heterogeneity and excluded variants showing significant heterogeneity with Q-SNP p-value < 5 × 10⁻⁸.

#### Multi-trait meta-analysis of psychiatric disorders

Next, we performed Multi-Trait Analysis of GWAS (MTAG)^61^ analysis to boost the power of the psychiatric disorder GWAS results using the shared pleiotropic loci with the correlated factors. MTAG takes the factor and psychiatric disorder GWAS as input and outputs the meta-analyzed results for each trait. To ensure that the results were reliable with low false positive rates, we calculated the maxFDR value, and SNPs with maxFDR values less than 0.05 were removed.

#### Identification of novel loci

After we obtained the MTAG summary statistics for each psychiatric trait, we assigned as genome-wide significantly associated SNPs those with p-value <5 × 10^−8^. We also identified the significant LD-independent loci using Plink^60^ clumping with p value threshold 5 × 10^−8^ and r^2^ < 0.1 within 3000kb region. To identify novel loci, we applied the same clumping thresholds to the original psychiatric disorder summary statistics to define existing loci, and considered non-overlapping loci from the multi-trait GWAS as novel. The significant loci were functionally annotated with the functional mapping and annotation (FUMA)^62^ platform using the European 1000 Genomes Project reference panel.

#### Gene prioritization and enrichment analyses

We used both positional and functional gene mapping methods to identify genes associated with the risk for each of the psychiatric disorders. We performed the positional gene mapping using MAGMA^63^ and applied Bonferroni correction to determine the significant genes. For functional gene mapping, we incorporated eQTL and chromatin interaction data through E-MAGMA^64^ and H-MAGMA^65^ analyses. E-MAGMA was performed using 13 brain tissue eQTL data from the GTEx consortium^66^ and H-MAGMA was performed using the brain tissue Hi-C data provided by the tool. In addition, we performed Transcriptome Wide Association Study (TWAS) and Summary data-based Mendelian Randomization with HEIDI (SMR&HEIDI)^67^ analyses. TWAS was performed using the Joint Tissue Imputation (TWAS_JTI)^68^ model with the S-PrediXcan^69^ tool using eQTL data from 49 GTEx v8 tissues. The aggregated Cauchy association test (ACAT) was performed to combine the p-values across different tissues. SMR analysis was performed using eQTL data from eQTLGen^70^, GTEx V8, and BrainMeta^71^. To ensure the robustness of the results, the heterogeneity in dependent instruments (HEIDI) test was applied, and genes with p<0.05 were subsequently excluded. The significant genes from all the gene mapping methods were identified with Bonferroni multiple testing correction. Finally, to identify novel associated genes, we compared our identified genes with those reported in the largest published psychiatric GWAS studies and in GWAS Catalog queries. Genes that had not been previously reported for the corresponding disorder were defined as novel.

Lastly, we performed gene set and tissue enrichment analyses to identify biological pathways and functional categories relevant to psychiatric disorders using MAGMA’s competitive gene-set test. The tested pathways and gene sets were from MsigDB^72^, and significant gene sets were selected after Bonferroni correction. Tissue enrichment analyses were performed using the GTEx V8 gene expression reference panel of 30 broad and 54 specific tissue types to evaluate whether associated genes were preferentially expressed in specific tissues and to provide biological context for the implicated loci.

## Data Availability

The data sources for the GWAS summary statistics included in this study are listed in Supplementary Table 1 for psychiatric disorders and Supplementary Table 2 for non-psychiatric disorders. The 23andMe GWAS summary statistics were accessed under a data use agreement through an approved application. The All of Us Research Program data were accessed through the All of Us Researcher Workbench under an approved researcher account. All other GWAS summary statistics analyzed in this study are publicly available from their original sources. The meta-analysis summary statistics generated in this study that exclude 23andMe data will be deposited in the GWAS Catalog and made publicly available upon publication. Meta-analysis and factor analysis summary statistics that include 23andMe data are subject to the 23andMe data use agreement and will be made available to qualified researchers through 23andMe in accordance with its data access policies.

## Acknowledgements

We would like to thank the research participants and employees of 23andMe Research Institute for making this work possible.

## Author contributions

QZ designed the study, performed the data analyses, and drafted and revised the manuscript. AT provided input in analysis, reviewed and edited the manuscript. PP conceived the study, supervised the research, and reviewed and edited the manuscript. All authors read and approved the final manuscript.

